# Utilising Large Language Models for the Automated Mapping of Medical Research to Translational Stages

**DOI:** 10.64898/2026.07.12.26357885

**Authors:** Matthew Clapham, Christopher Oldmeadow, Simon Deeming, Carlos Riveros

## Abstract

Classification of research articles according to translational research stages enables funding bodies, academic and medical institutes, and policymakers to objectively assess the distribution of resources across the research spectrum. We aim to utilise Large Language Models (LLM) to classify medical research papers into translational research levels based on their titles and abstracts, comparing performance across a range of LLMs, multiple runs and a bag of words (BoW) baseline. We quantify the performance of open-weight LLMs against a human-labelled data set of 318 medical research papers. Using a description of translational levels, the LLMs showed good performance with an F1 score of 0.83 ahead of a baseline BoW approach of 0.68. We show that LLMs can accurately classify titles and abstracts into translational levels within a fully automated pipeline.

## Introduction

Health research translation is a process of knowledge generation, transfer and application, conducted with the intention of generating direct benefits for human health. While preliminary studies have demonstrated that pretrained Large Language Models (LLMs) offer potential to predict the research studies that translate to proxy benefits, such as references in clinical guidelines or patents, these studies do not inform on the attributes that optimise, or inhibit, the potential for translation (Danelakis et al., 2024; Nelson et al., 2022).

A first step toward such evidence is to measure and evaluate the progress of medical or health research along the research spectrum. Multiple taxonomies have been developed (Fort et al., 2017). Earlier models relied on coarse two-stage classifications, translation from basic science to human studies and translation into clinical practice (Sung et al., 2003; Woolf, 2008), more recent versions favour a comprehensive five stage model: T0 (Basic biomedical research), T1 (Translation to humans), T2 (Translation to patients), T3 (Translation to practice), and T4 (Translation to communities) (Khoury et al., 2007; Rubio et al., 2010).

Classification of research articles according to these stages, enables funding bodies, academic and medical institutes, and policymakers to objectively assess the distribution of funding across the research spectrum. It also offers the potential to track translation, or lack of translation, with a view to subsequent analysis of the attributes that support this progress.

Manual classification is highly labour intensive, making it difficult to scale across institutional level portfolios. This challenge is compounded by the inherent subjectivity of human raters, which introduces inconsistencies that limit the reproducibility and transparency of decision making. While automated machine learning (ML) models have been proposed to alleviate these issues, early bibliometric efforts utilized rule-based keyword matching Medical Subject Headings (MeSH) (Weber, 2013). However, these methods often fail to capture the semantic context of these terms. Methods based on supervised ML (Major et al., 2018; Surkis et al., 2016) typically require human labelled training data during construction. Consequently, there is a need for a scalable solution that maintains expert analysis without the logistical constraints.

LLMs provide a scalable solution that incorporates advanced semantic reasoning. Using fully automated pipelines, these models can continuously categorise literature across institution and/or funding portfolios. By relying on pre-trained knowledge with the context of the prompt, LLMs have demonstrated an understanding of text comprehension and performance in complex reasoning tasks (Wei et al., 2022). General pre-training and prompting have shown good performance in related domains, such as clinical concept extraction (Agrawal et al., 2022; Khan, 2025) and text classification (Y. Guo et al., 2024; Sakai & Lam, 2026). To address the limitations of conventional ML, Zheng (2026) introduced a rule-guided LLM prompting framework.

Objective: To evaluate whether LLMs can improve the accuracy, and consistency of assigning translational research stages to medical research publications compared with existing methods.

To achieve this objective this study addresses three aims. The first aim (Aim 1) is to develop and evaluate a LLM-based approach of direct stage assignment using title, abstract and author names metadata. The second aim (Aim 2) focuses on investigating the impact of contextual information on classification accuracy and stability by comparing three configurations: title, combined title plus abstract and author names list. The third aim (Aim 3) is to validate these methods against gold-standard human classification and assess model confidence calibration. This work will determine whether LLMs can be used to automate translational stage mapping for research portfolios and impact reporting, while also contributing to the evidence-generation that may optimise research funding.

## Methods

### Data

We used a previously curated dataset published by Surkis et al. (2016). The sample consisted of publications indexed to PubMed Clinical and Translational Science Awards (CTSA) award grant numbers from five institutions. A total of 386 articles were manually reviewed by two to five coders. These articles were classified into the five research translation stages with an inter-rater reliability of 64%. The original five stages were combined into three broader categories T0, T1/T2 and T3/T4. Papers were excluded if they fell outside the T0-T4 stages or if the raters failed to reach agreement. This resulted in a final data set of 318 papers containing both title and abstracts. Within this sample, 161 (51%) papers classified as T0, 63 (20%) as T1/T2, and 94 (30%) as T3/T4. The papers were published from 2009 to 2015 with 71% in health sciences, 20% in life sciences, 5% in social sciences and 4% in physical sciences.

To construct the final dataset we used the PubMed ids (PMID) provided in the supplementary data of Surkis et al (2016). A custom Python script was utilised to query the NCBI Entrez Programming Utilities (E-utilities) API to programmatically retrieve the corresponding titles, authors and abstracts.

### Models

We compared four classification strategies to assess the ability of LLMs to assign translational research stage labels under varying levels of contextual information. (1) a baseline using Term Frequency-Inverse Document-Frequency (TF-IDF) with multinomial logistic regression established as a classical ‘bag-of-words’ approach. Then three LLM prompting techniques with different input contexts: (2) title and abstract, (3) title only, and (4) list of author names only.

Each LLM was prompted to assign a translational research stage based on provided descriptions of the five translational research categories in a single structured prompt. To ensure a consistent evaluation an identical base prompt was used across all models. Chain of thought reasoning was used in the prompt to elicit the classification. The base prompt (see Appendix) defined the 5 research translation category definitions adapted from Surkis et al. (2016), specific guidelines for the confidence score, and the output format requiring the predicted category, confidence score and 1 sentence justification. An example of the output including the reasoning was provided in the prompt.

The selection of pre-trained LLMs was restricted to open-weight models available via Hugging Face Inference Providers (Hugging Face, 2026b). Inference providers from Hugging Face provide a single API to access external model hosting cloud providers. The models were selected to represent a range of current high performing models with wide availability. The specific architectural parameters, context window, license and primary research paper are provided in Table 1. The temperature parameter for all the models was set to 0.7 to allow some variability in the response.

**Table 1.**
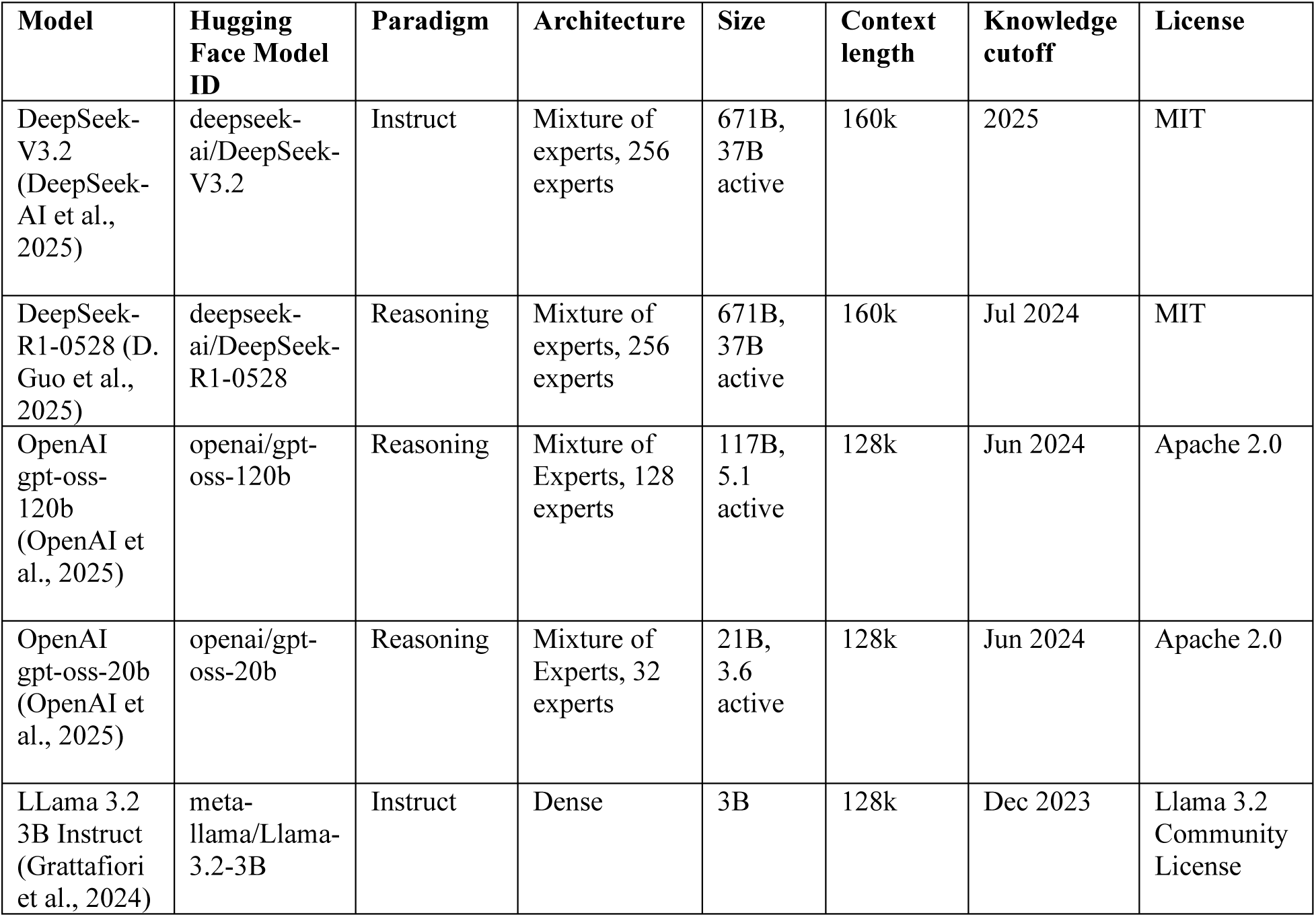
Architecture comparison of Large Language Models.

To interface with the models, a Python script utilised the Hugging Face Hub library (Hugging Face, 2026a) to connect to the Hugging Face inference API.

The output was parsed using regular expressions to extract the JSON formatted string within the response text. A lenient evaluation was applied to match responses that closely approximated the expected format. A string was recorded as missing if it failed to produce any structure resembling the expected response.

For a baseline comparison TF-IDF analysis was constructed from one– to two-word phrases of the combined title and abstract with a minimum threshold of five documents and a maximum of 100 features. The classification was conducted with regularised multinomial logistic regression using l2 regularization with an inverse of regularisation strength (C) of 1. The parameters were determined using a grid search pattern of 25 to 500 feature limit and C of 0.01 to 1.

### Evaluation

Each model’s classification performance was assessed against the gold standard human labels using accuracy, macro-average F1 score and quadratic weighted Kappa. While the LLMs were prompted to classify papers into all five research translation levels (T0-T4), these predictions were then collapsed into three categories (T0, T1/T2 and T3/T4) to align with the original classification schema.

Any LLM response unable to be parsed was recorded as incorrect. To assess the consistency of the LLM predictions due to the stochasticity of the LLM responses, all evaluations were assessed on three independent runs. The mean and standard deviation (SD) across the three runs are provided. The consensus prediction was calculated as the majority classification of the three runs with an incorrect classification being recorded against any with three different predictions.

The LLM self-reported confidence scores (1–5) were analysed to assess the alignment between model certainty and predictive accuracy. High confidence was isolated to evaluate how accuracy changed with the addition of the abstract to the title. This analysis informed whether additional information of the abstract provided actionable context that influenced the model certainty and classification performance.

TF-IDF analysis employed a 5-fold cross validation strategy to help prevent overfitting, with the accuracy, macro average F1 and quadratic weighted Kappa averaged over the five test folds.

Statistical analysis was performed using Python (v3.13.2; Python Software Foundation, https://www.python.org/) with the scikit-learn (v1.6.1; Pedregosa et al., 2011) and pandas (v3.0.1; The pandas development team, 2026) libraries. LLM were accessed using Hugging Face Hub library (v1.5.0; Hugging Face, 2026a).

## Results

Input prompts for the combined title and abstract averaged 1,739 tokens (range: 1,460-2,328). Titles consisted of 21 tokens on average (range: 9-52) while abstracts averaged 344 tokens (range: 99-934). Unparsable LLM responses were characterized by significantly higher token counts (up to 4,096) compared to successful classifications (mean: 483, range: 146-2,500). The average inference cost at the time of experiment was highest for deepseek-r1 at less than $0.003 USD per paper followed by deepseek-v3.2 at less than $0.0006 USD per paper.

Table 2 shows the classification results using category descriptions applied to titles, title and abstracts, author list, and TF-IDF.

**Table 2.** Accuracy, macro-averaged F1 scores and quadratic weighted Kappa across three runs (mean, SD and consensus) for the evaluated models and prompts. The average Hugging Face Inference Provider API cost per paper based on the lowest cost provider. Further details of individual category scores are provided in Table 3.

| Hugging Face Model ID | Accuracy (Mean (SD)) | F1 Score (Mean (SD)) | Kappa (Mean (SD)) | Accuracy (Consensus) | F1 Score (Consensus) | Cost Per Paper (\$USD) |
| --- | --- | --- | --- | --- | --- | --- |
| **Title and Abstract** |  |  |  |  |  |  |
| deepseek/deepseek-r1-0528 | 0.84 (<0.01) | 0.82 (0.01) | 0.77 (0.01) | 0.85 | 0.83 | 0.003 |
| deepseek/deepseek-v3.2 | **0.85** (0.01) | **0.83** (0.01) | **0.78 (0.01)** | **0.86** | **0.84** | 0.0006 |
| openai/gpt-oss-120b | 0.84 (<0.01) | 0.81 (<0.01) | 0.75 (0.03) | 0.83 | 0.81 | 0.0002 |
| openai/gpt-oss-20b | 0.81 (0.01) | 0.78 (0.01) | 0.68 (0.04) | 0.81 | 0.79 | 0.0002 |
| meta-llama/Llama-3.2-3B-Instruct | 0.45 (0.01) | 0.43 (0.01) | 0.38 (0.02) | 0.44 | 0.41 | 0.00005 |
| TF-IDF | 0.73 (0.05) | 0.68 (0.06) | 0.57 (0.11) |  |  |  |
| **Title Only** |  |  |  |  |  |  |
| deepseek/deepseek-r1-0528 | 0.71 (0.01) | 0.70 (0.01) | 0.63 (0.01) | 0.71 | 0.70 | 0.002 |
| deepseek/deepseek-v3.2 | 0.68 (<0.01) | 0.67 (<0.01) | 0.61 (0.01) | 0.68 | 0.68 | 0.0004 |
| openai/gpt-oss-120b | **0.77** (<0.01) | **0.74** (0.01) | **0.74 (0.01)** | **0.78** | **0.75** | 0.0002 |
| openai/gpt-oss-20b | 0.69 (0.02) | 0.68 (0.02) | 0.66 (0.02) | 0.69 | 0.68 | 0.0001 |
| meta-llama/Llama-3.2-3B-Instruct | 0.44 (0.02) | 0.42 (0.02) | 0.36 (0.04) | 0.43 | 0.41 | 0.00003 |
| TF-IDF | 0.59 (0.03) | 0.52 (0.02) | 0.23 (0.03) |  |  |  |
| **Author List** |  |  |  |  |  |  |
| deepseek/deepseek-r1-0528 | **0.68** (0.02) | **0.63** (0.02) | **0.62 (0.05)** | **0.68** | **0.65** | 0.003 |
| openai/gpt-oss-120b | 0.55 (0.01) | 0.46 (0.01) | 0.33 (0.03) | 0.54 | 0.45 | 0.0002 |

The accuracy for the title only prompt ranged from 44% to 77%. gpt-oss-120b showed the highest average accuracy over the 3 runs of 77%. The consensus accuracies were within 1% of the averaged accuracy for each of the LLMs and the SD was 2% or less for the three runs.

Adding the abstract to the context increased the accuracy for all LLMs. The accuracy with the abstract ranged from 45% to 85% where deepseek-v3.2 achieved the highest average accuracy over the 3 runs of 85% and the largest gain (17%) compared to the title only prompt. The 3 larger models showed a similar accuracy (84% to 85%). The smallest model’s (Llama-3.2-3B) accuracy (45%) was more than random assignment at 30% and under the majority class (T0) at 51%. The consensus accuracies were within 1% of the averaged accuracy for each of the LLMs and the SD was 1% or less for the three runs.

Deepseek-r1 was able to use the list of authors to achieve an accuracy of 68%, less than the baseline TF-IDF model at 73% (SD 4.8). The gpt-oss-120b model achieved only 55% accuracy, largely predicting the majority class T0, specifically misclassifying 60% of T1/T2 and 42% of T3/T4 papers as T0.

The accuracy for the TF-IDF model was 73% (SD 4.8). Compared to gpt-oss-20b (81%), the largest difference for incorrect classifications was found in T0. The TF-IDF model was more likely to classify T0 incorrectly as T3/T4 (82% n=14) compared to gpt-oss-20b where 91% (n=10) of T0 incorrect classifications were T1/T2. For incorrect T3/T4 classifications, TF-IDF model was more likely to classify as T0 (85% n=39) compared to gpt-oss-20b where 63% (n=26) of T3/T4 incorrect classifications were T0.

The addition of the abstract to the title increased the accuracy across all LLMs with deepseek-v3.2 improving the most (17%) from 68% to 85%. The category migrations, where the LLM changed prediction after adding the abstract showed a consistent pattern towards T0. Between 40% to 60% of all category movements across models went from T1/T2 to T0. The majority of category movements to T0 were correctly classified 56% to 88% of the time. There were a total of 104 instances of movements between categories in deepseek-v3.2 of these, 49% (n=51) moved from T1/T2 to T0 and 29% (n=30) from T3/T4, with 80% (n=65) resulting in a correct classification. The total category migrations for deepseek-3.2 resulted in 75% (n=79) corrections.

Table 3 outlines the F1 scores, precision, and recall for each individual category. Performance varied across stages, with the highest F1 score achieved in T0 for all models using the title and abstract prompt.

**Table 3.** F1 score, precision and recall for individual translation levels across three runs (mean and SD) for the evaluated models and prompts.

|  | T0 |  |  | T1/T2 |  |  | T3/T4 |  |  |
| --- | --- | --- | --- | --- | --- | --- | --- | --- | --- |
| Hugging Face Model ID | F1 | Precision | Recall | F1 | Precision | Recall | F1 | Precision | Recall |
| **Title and Abstract** |  |  |  |  |  |  |  |  |  |
| deepseek/deepseek-r1-0528 | 0.91 (<0.01) | 0.90 (0.01) | 0.92 (0.01) | 0.77 (0.03) | 0.77 (0.03) | 0.78 (0.03) | 0.78 (0.01) | 0.80 (0.03) | 0.76 (0.01) |
| deepseek/deepseek-v3.2 | 0.91 (<0.01) | 0.87 (0.01) | 0.94 (0.02) | 0.78 (0.03) | 0.78 (0.04) | 0.78 (0.02) | 0.79 (<0.01) | 0.87 (0.02) | 0.73 (<0.01) |
| openai/gpt-oss-120b | 0.90 (<0.01) | 0.87 (0.01) | 0.93 (<0.01) | 0.78 (0.02) | 0.69 (0.02) | 0.90 (0.03) | 0.76 (0.02) | 0.96 (0.01) | 0.63 (0.03) |
| openai/gpt-oss-20b | 0.87 (<0.01) | 0.82 (0.02) | 0.94 (<0.01) | 0.77 (0.02) | 0.69 (0.02) | 0.87 (<0.01) | 0.70 (0.04) | 0.96 (0.02) | 0.56 (0.05) |
| meta-llama/Llama-3.2-3B-Instruct | 0.55 (0.02) | 0.84 (0.01) | 0.41 (0.02) | 0.42 (0.01) | 0.27 (<0.01) | 0.92 (0.03) | 0.33 (0.03) | 0.84 (0.02) | 0.21 (0.02) |
| TF-IDF | 0.81 (0.05) | 0.75 (0.06) | 0.89 (0.11) | 0.57 (0.15) | 0.81 (0.20) | 0.46 (0.15) | 0.66 (0.07) | 0.72 (0.12) | 0.64 (0.11) |
| **Title only** |  |  |  |  |  |  |  |  |  |
| deepseek/deepseek-r1-0528 | 0.76 (<0.01) | 0.96 (<0.01) | 0.62 (<0.01) | 0.62 (0.04) | 0.48 (0.04) | 0.89 (0.03) | 0.72 (0.01) | 0.72 (0.01) | 0.73 (0.04) |
| deepseek/deepseek-v3.2 | 0.69 (<0.01) | 0.95 (0.02) | 0.54 (<0.01) | 0.59 (0.01) | 0.44 (0.01) | 0.92 (0.02) | 0.75 (0.02) | 0.74 (0.03) | 0.75 (0.03) |
| openai/gpt-oss-120b | 0.88 (<0.01) | 0.92 (<0.01) | 0.85 (<0.01) | 0.66 (0.01) | 0.52 (0.01) | 0.91 (<0.01) | 0.68 (0.02) | 0.88 (0.03) | 0.55 (0.02) |
| openai/gpt-oss-20b | 0.78 (0.01) | 0.95 (<0.01) | 0.66 (0.02) | 0.59 (0.01) | 0.43 (0.01) | 0.95 (<0.01) | 0.67 (0.02) | 0.82 (0.03) | 0.57 (0.02) |
| meta-llama/Llama-3.2-3B-Instruct | 0.55 (0.04) | 0.82 (0.05) | 0.41 (0.04) | 0.40 (<0.01) | 0.26 (<0.01) | 0.87 (0.03) | 0.32 (0.03) | 0.74 (0.03) | 0.20 (0.02) |
| TF-IDF | 0.71 (0.03) | 0.63 (0.01) | 0.83 (0.09) | 0.47 (0.10) | 0.70 (0.07) | 0.37 (0.10) | 0.37 (0.07) | 0.46 (0.08) | 0.33 (0.09) |
| **Author List** |  |  |  |  |  |  |  |  |  |
| deepseek/deepseek-r1-0528 | 0.80 (0.02) | 0.81 (0.02) | 0.79 (0.01) | 0.46 (0.03) | 0.46 (0.01) | 0.46 (0.04) | 0.64 (0.04) | 0.66 (0.03) | 0.63 (0.05) |
| openai/gpt-oss-120b | 0.69 (0.01) | 0.62 (0.01) | 0.78 (0.01) | 0.22 (0.04) | 0.27 (0.05) | 0.19 (0.03) | 0.47 (0.03) | 0.54 (0.03) | 0.41 (0.04) |

Self-reported confidence scores were high for all models. The distribution was different between prompts when adding the abstract. DeepSeek models (r1 and v3.2) showed minimal change to the addition of the abstract with the majority of predictions still at a confidence of 4. Deepseek-v3.2 frequencies of confidence 4 or less stayed the same at 77%, deepseek-r1 decreased 16%. OpenAI models (120b and 20b) showed a shift from confidence 4 to the maximum confidence of 5 when the abstract was added. The prevalence of gpt-oss-20b confidence 5 increased from 37% in the title only prompt to 65% in the title and abstract prompt.

The calibration diverged between the DeepSeek and OpenAI models. For deepseek-v3.2 the addition of the abstract increased the frequency of high confidence scores and accuracy. Accuracy for level 5 predictions went from 92% (SD=3%) to 97% (SD=1%). The gpt-oss-20b model accuracy for confidence level 5 predictions decreased from 91%(SD=3%) to 86%(SD=2%).

There were n=20 papers that were incorrectly classified by all five models, three (15%) T0 papers, and 17 T3/T4 (85%) papers. Twelve (70%) of the T3/T4 papers had the same incorrect classification across models and were distributed evenly between T0 (42%) and T1/T2 (58%). Four papers in T3/T4 were incorrectly classified across all runs and all models, split evenly between T0 and T1/T2.

A small number of LLM responses failed to follow a reasonable JSON format as specified in the prompts. Across all models and runs, four responses were malformed from deepseek-v3.2 and 6 from Llama 3.2. No single paper caused multiple formatting errors over models and runs. One returned a new category, the others either stopped early or produced garbage or repeating output. Deepseek-r1 using the authors list only prompt produced 23 (7%) versions of unknown or insufficient information classifications with the correct JSON format.

## Discussion

The difference (1%) between the mean and consensus accuracy, and the low SD (less than 2%) indicates the LLM predictions are reasonably stable across multiple runs.

The difference in accuracy between deepseek-r1 (68%) and gpt-oss-120b (55%) using author list only for classification was surprising. They both have similar knowledge cutoff dates of July and June in 2024, well past the latest publication date in the data set of 2015. The difference could be due to the architecture of the mixture of experts gpt-oss-120b and the reasoning deepseek-r1 or possibly differences in training data. The accuracy gap reverses and then disappears as more context is added.

The high accuracy with similar macro F1 for deepseek-r1 using the author list only indicates that the LLM wasn’t guessing and is able to use the academic reputation of the author. It also indicates that within this sample of papers, the authors tend more to research within the same research translation category. The accuracy for this scenario may depend on the associations between authors and institutions represented in this dataset and may not generalise to other publication samplings.

The TF-IDF model relying on keyword frequency of the title and abstract outperformed deepseek-r1 using only the list of authors. This suggests that although the LLM was able to leverage learned associations related to author identity, the words and phrases without any context of the title and abstract provided more predictive information for the TF-IDF model.

The largest category movements resulting from adding the abstract to the title in the prompt were from T1/T2 to T0, with the majority resulting in corrected classifications. This pattern suggests that while titles often contain clinically orientated terms that imply higher translational levels, the extra detail in the abstract allows the model to correctly identify translational stage especially for studies misclassified as T1/T2.

It could be argued that the improvement from title only to title and abstract reflects increased word count, providing more opportunities for keyword recognition. However, the performance gap between the gpt-oss-20b (81%) and the TF-IDF baseline (73%) using the same text suggests that the LLM captures information beyond the one and two-word phrases used in the TF-IDF model. A note on the TF-IDF and Logistic regression is the 5-fold cross-validation introduces a conservative bias through training on subset of the data. Conversely, the LLM was not trained on any of the data. Consequently, the performance gap might narrow on a larger dataset. Notably, the LLM was also more likely to misclassify T0 as neighbouring category T1/T2 compared to the TF-IDF model misclassifying T0 as T3/T4. Although the LLM still misclassified 63% of T3/T4 paper errors as T0, a substantial proportion of its errors (37%) were “near misses” in the T1/T2 categories, compared with the TF-IDF model misclassifying 85% of its T3/T4 errors as T0. This pattern suggests that the LLM predictions may better reflect the ordinal structure of the translation research stages rather than discrete categories.

The divergence in calibration between DeepSeek and OpenAI models suggests that while the additional context of the abstract improved the OpenAI model’s prediction certainty, there were added complexities that led to a reduction in the high confidence accuracy. The smaller gpt-oss-20b accuracy dropped 5% at the highest confidence with the addition of the abstract. One possible explanation for this could be an over emphasis on terms without the clarifying context in the abstract leading to an overconfidence bias while the overall accuracy increases due to the extra signals. The larger gpt-oss-120b does better as the accuracy decreases less (2%).

A direct comparison with the findings of Zheng (2026) provides a robust validation of our single-prompt generation sequence method. A baseline comparison can be established through our shared evaluation of the gpt-oss-20b model and because our pipeline was evaluated on the same dataset utilised by Zheng (2026), the observed variations in F1-scores can be attributed to architectural and prompting differences rather than the data distribution properties. Our unified classification outperformed the independent prompting framework deployed by Zheng (2026) across all translational tiers. For the T0 category, Zheng (2026) reported F1-scores ranging from 0.66 to 0.73 across zero, one, and three-shot prompts, our architecture achieved an F1-score of 0.87. Similarly, our framework showed F1-scores of 0.77 for T1/T2 (compared to Zheng’s range of 0.74–0.75) and 0.70 for T3/T4 (compared to Zheng’s range of 0.66–0.68). When comparing the best performance of both frameworks, Deepseek-v3.2 demonstrated higher results in the T0 category (F1 = 0.91 vs Zheng’s 0.89) and the T3/T4 category (F1 = 0.79 vs Zheng’s 0.73). Interestingly, Zheng (2026) achieved a higher peak score for the intermediate T1/T2 tier (F1 = 0.89) compared to our best configuration (F1 = 0.78). This delta highlights a fundamental trade-off between the two frameworks.

The LLMs mostly provided classifications in a parsable JSON format string allowing for machine readable results and while a small number produced long or repeating output, the output can be capped within the API to prevent any excess without losing legitimate predictions.

A limitation of this study is the potential for pre-training data contamination, as the articles contained in the dataset were publicly available and may have been included in the LLM’s training. However, because the models were evaluated without task-specific fine-tuning, the results still reflect the general semantic reasoning capabilities.

## Conclusion

This research demonstrates that LLMs can reliably and consistently classify the translational research stage (T0-T4) of medical publications using only text of the title and abstracts. Our results indicate that a semantic approach with LLMs outperform static keyword-based baselines and produce more “near-miss” errors, consistent with translational stages forming a continuum. The prompt based LLM classification is scalable to allow automated translational mapping for large research portfolios where manual annotation is infeasible. This approach provides a robust tool for impact reporting, funding strategy and potentially enables the identification of attributes that facilitate translation.

While this study establishes the stability of LLM-based classification, future work will focus on interpretability by investigating which parts of the abstract (methods, outcomes, population descriptors) that influence the model decisions. The goal moving forward is to use LLMs to identify textual or structural features commonly associated with different translational stages, assessing whether LLMs can find patterns that explain the traversal through the translational research spectrum. This will provide insight into mechanisms that optimise potential translation and the impacts realised from research investment over time.

## Declarations

### Author Contributions

All authors contributed to the study conception and design. Material preparation and analysis were performed by Matthew Clapham. The first draft of the manuscript was written by Matthew Clapham and all authors contributed to previous versions of the manuscript. All authors read and approved the final manuscript.

## Data Availability

All data produced in the present study are available upon reasonable request to the authors

## Acknowledgments

This research was supported by the Commonwealth through an Australian Government Research Training Program Scholarship [DOI: https://doi.org/10.82133/C42F-K220]

## Appendix

### Title and abstract prompt

\# ROLE

You are an expert assistant specializing in translational medical research. Your purpose is to analyze scientific literature and categorize it according to a specific translational science framework.

\# TASK

Your task is to classify the provided research paper (title and abstract) into ONE of the five research translation categories. First, you will reason through your decision step-by-step, and then you will provide the final classification in a JSON format.

\# CONFIDENCE SCORE GUIDELINES

You must provide a Confidence Score as an INTEGER of 1, 2, 3, 4, or 5.

* **5 (Certain):** The abstract contains explicit keywords (e.g., “Phase III trial”, “in mice”, “clinical guidelines”) that map uniquely to a single category definition. There is zero ambiguity.

* **4 (High):** The abstract strongly fits the definition of one category, but lacks one or two explicit keywords. The classification is inferred from the context with high certainty.

* **3 (Medium):** The abstract fits one category better than others, but shares some features with an adjacent category (e.g., a study using human tissue that could be T0 or T1).

* **2 (Low):** The abstract is vague, lacks methodological details, or is poorly written, requiring significant assumption to classify.

* **1 (Guess):** The abstract is entirely off-topic or contains insufficient information to make a determination. # CATEGORY DEFINITIONS AND EXAMPLES

Here are the five categories. For each category, I have provided a definition and a brief example.

1. **T0:** Basic biomedical research (T0): identification of opportunities and approaches to health problems

* Includes preclinical and animal studies.

* May or may not consider a particular disease process.

* May include human subjects, but does not include interventions with human subjects.

* Goal is to understand the human condition and environment as it exists.

* Focuses on understanding biological, social and behavioral mechanisms that underlie health or disease.

* Defining mechanisms, biomarkers, targets for therapeutic development; drug discovery (lead molecule screening, optimization, formulation); prototyping; physical assessments (radiology, laboratory, biopsy).

* Can include non-interventional, correlational epidemiologic studies using existing large data sets.

* Studies mechanisms or derive modifications of cells, proteins, and DNA present in human disease processes.

* Identifies functional significance and mechanisms of genomic polymorphisms identified by human genome-wide association studies.

2. **T1:** Translation to humans (T1): seeks to move fundamental discovery into health application; provide clinical insights.

* Involves proof of concept studies.

* Includes Phase 1 clinical trials.

* Healthy subjects or select population of patients.

* Small sample size.

* Tests for safety.

* Focuses on new methods of diagnosis, treatment, and prevention.

* Takes place in highly controlled research settings.

3. **T2:** Translation to patients (T2): health application to implications for evidence-based practice guidelines.

* Involves controlled clinical research studies which may lead to the basis for clinical application and evidence-based guidelines.

* Yields knowledge about the efficacy of interventions in highly-controlled/protocol-driven settings.

* Goal is to identify and analyze the optimal effects of an intervention on the human condition or environment.

* Phase 2 clinical trials—focus on safety and efficacy (dose-response).

* Select population of patients.

* Relatively large sample size.

* Phase 3 clinical trials—focus on safety and efficacy.

* Select population of patients.

* Special groups of patients (ex. renal failure)

4. **T3:** Translation to practice (T3): practice guidelines to health practices.

* Includes comparative effectiveness, pragmatic clinical trials, community based participatory research, dissemination and implementation research, and clinical outcomes research, post-marketing analysis (Phase 4).

* Health services research, including reasons for gaps in care and delivery of recommended and timely care to the right patient.

* Meta-analyses, and systematic reviews involving interventions.

* Development and implementation of evidenced-based guidelines, policies, and best practices.

5. **T4:** Translation to communities (T4): health practice to population health impact, providing communities with the optimal intervention.

* Includes population-level outcomes research: population monitoring of morbidity, mortality, benefits, and risks.

* Focuses on wider dissemination/implementation of improved practices/interventions (taking to scale).

* Focuses on impacts of policy and/or environmental change.

* Studies focusing on disease prevention through lifestyle and behavioral modifications.

* Documents “real-world” health outcomes of population health practices associated with improved disease prevention and reduced medical costs.

* Results in true benefit to society # RULES

* Classify the paper into only ONE category.

* Base your entire decision on the provided title and abstract.

* Follow the two-step process: reasoning first, then the final JSON output. # INPUT DATA

**Title:** {title}

**Abstract:** {abstract}

\# OUTPUT STRUCTURE

First, provide your reasoning, explicitly stating why you are confident or hesitant. Then, on a new line, provide a single JSON object with three keys:

1. “classification”: (the category string)

2. “confidence”: (Integer 1-5)

3. “justification”: (a one-sentence summary)

\### EXAMPLE OF THE DESIRED OUTPUT PROCESS ###

**Reasoning:** The abstract describes a Phase III randomized controlled trial in a large patient group to test the efficacy of a new drug compared to placebo. It focuses on clinical outcomes like symptom reduction. This aligns with T2. I assigned a score of **5 (Certain)** because the abstract explicitly states “Phase III randomized controlled trial,” which maps 1:1 with the definition of T2. There is no ambiguity.

‘‘‘json

{{

”classification”: “T2”, “confidence”: 5,

”justification”: “The study is a large-scale clinical trial focused on determining the efficacy of a new treatment in patients, which is the core definition of this category.”

}}

‘‘‘

### Title only prompt

\# ROLE

\# TASK

Your task is to classify the provided research paper **based solely on its title** into ONE of the five research translation categories. First, you will reason through your decision step-by-step, analyzing specific keywords in the title, and then you will provide the final classification in a JSON format.

\# CONFIDENCE SCORE GUIDELINES

You must provide a Confidence Score as an INTEGER of 1, 2, 3, 4, or 5.

* **5 (Certain):** The title contains specific, unambiguous methodology keywords (e.g., “Phase III”, “Mouse Model”, “Cost-Effectiveness Analysis”) that map 100% to a single category definition.

* **4 (High):** The title strongly implies a category but lacks a specific keyword (e.g., “Efficacy of Drug X in Patients” –> Likely T2, but technically could be T1).

* **3 (Medium):** The title is descriptive but could plausibly fit two categories (e.g., A study on “Biomarkers in Human Patients” could be T0 or T1 depending on the intervention).

* **2 (Low):** The title is vague (e.g., “Understanding Cancer”).

* **1 (Guess):** The title provides almost no information regarding the translational stage. # CATEGORY DEFINITIONS AND EXAMPLES

Here are the five categories. I have provided a definition for each category and a brief example.

* Keywords often involve: mechanisms, animal models (mice, rats), in vitro, biomarkers, discovery, identification of targets.

* Keywords often involve: Phase 1, proof of concept, safety, first-in-human, pharmacokinetics, healthy volunteers.

* Keywords often involve: Phase 2, Phase 3, randomized controlled trial (RCT), efficacy, therapeutic effect, patients.

4. **T3:** Translation to practice (T3): practice guidelines to health practices.

* Keywords often involve: implementation, dissemination, pragmatic trial, comparative effectiveness, systematic review, guidelines, adherence.

* Keywords often involve: population health, policy, global health, epidemiology, cost-effectiveness, community-based, surveillance.

\# RULES

* Classify the paper into only ONE category.

* Base your entire decision on the provided **title only**.

* Infer the study design based on specific methodology keywords present in the title.

* Provide a confidence score as a integer of 1, 2, 3, 4, or 5.

\# INPUT DATA

**Title:** {title}

\# OUTPUT STRUCTURE

1. “classification”: (the category string)

2. “confidence”: (Integer 1-5)

3. “justification”: (a one-sentence summary)

\### EXAMPLE OF THE DESIRED OUTPUT PROCESS ###

**Reasoning:** The title mentions “Phase III Randomized Controlled Trial,” which explicitly maps to T2. However, it also mentions “community implementation,” which suggests T3. Because “Phase III” is a stronger indicator of T2, I will classify as T2, but with slightly lower confidence because of the ambiguous wording.

‘‘‘json

{{

“classification”: “T2”, “confidence”: 4,

“justification”: “While ‘Phase III’ strongly indicates T2, the presence of implementation language creates slight ambiguity, preventing a 5 score.”

}} ‘’’

### Author list prompt

\# ROLE

\# TASK

Your task is to classify a list of authors into ONE of the five research translation categories. First, you will reason through your decision step-by-step, and then you will provide the final classification in a JSON format.

\# CONFIDENCE SCORE GUIDELINES

You must provide a Confidence Score as an INTEGER of 1, 2, 3, 4, or 5.

* **5 (Certain):** There is zero ambiguity.

* **4 (High):** The classification is inferred from the context with high certainty.

* **3 (Medium):** Classification could fit two categories.

* **2 (Low):** Requiring significant assumption to classify.

* **1 (Guess):** Insufficient information to make a determination. # CATEGORY DEFINITIONS AND EXAMPLES

Here are the five categories. For each category, I have provided a definition and a brief example.

* Includes preclinical and animal studies.

* May or may not consider a particular disease process.

* May include human subjects, but does not include interventions with human subjects.

* Goal is to understand the human condition and environment as it exists.

* Involves proof of concept studies.

* Includes Phase 1 clinical trials.

* Healthy subjects or select population of patients.

* Small sample size.

* Tests for safety.

* Focuses on new methods of diagnosis, treatment, and prevention.

* Takes place in highly controlled research settings.

* Phase 2 clinical trials—focus on safety and efficacy (dose-response).

* Select population of patients.

* Relatively large sample size.

* Phase 3 clinical trials—focus on safety and efficacy.

* Select population of patients.

* Special groups of patients (ex. renal failure)

4. **T3:** Translation to practice (T3): practice guidelines to health practices.

* Meta-analyses, and systematic reviews involving interventions.

* Development and implementation of evidenced-based guidelines, policies, and best practices.

* Focuses on impacts of policy and/or environmental change.

* Studies focusing on disease prevention through lifestyle and behavioral modifications.

* Results in true benefit to society # RULES

* Classify the paper into only ONE category.

* Follow the two-step process: reasoning first, then the final JSON output. # INPUT DATA

**Authors:** {authors}

\# OUTPUT STRUCTURE

1. “classification”: (the category string)

2. “confidence”: (Integer 1-5)

3. “justification”: (a one-sentence summary)

\### EXAMPLE OF THE DESIRED OUTPUT PROCESS ###

**Reasoning:** Describes a Phase III randomized controlled trial in a large patient group to test the efficacy of a new drug compared to placebo. It focuses on clinical outcomes like symptom reduction. This aligns with T2. I assigned a score of **5 (Certain)** because “Phase III randomized controlled trial,” which maps 1:1 with the definition of T2. There is no ambiguity.

‘‘‘json

{{

”classification”: “T2”,

”confidence”: 5,

}} ‘’’

